# Predictors and barriers to vaccination among older Syrian refugees in Lebanon: a multi-wave survey

**DOI:** 10.1101/2022.12.15.22282964

**Authors:** Berthe Abi Zeid, Tanya El Khoury, Hala Ghattas, Marwan F. Alawieh, Zeinab Ramadan, Sirine Anouti, Sawsan Abdulrahim, Stephen J. McCall

**Author notes:** Senior authors /Corresponding authors. Joint first authors (Berthe Abi Zeid, Tanya El Khoury).

## Abstract

**Background:** Access to vaccination is important to prevent morbidity and mortality due to COVID-19 among older Syrian refugees. This study aimed to develop an internally validated predictive model for COVID-19 vaccination amongst older Syrian refugees in Lebanon and understand barriers to vaccination.

**Methods:** This multi-wave longitudinal study was conducted through telephone interviews between September 2020 and March 2022 in Lebanon. Syrian refugees aged 50 years or older were invited to participate from a list of households that received assistance from a humanitarian organization. The outcome was self-reported COVID-19 vaccination status. Logistic regression was used to identify predictors of vaccination uptake. Validation was completed internally using bootstrapping methods.

**Findings:** Out of 2,906 participants (median[IQR] age:58[55-64]; 52.9% males), 1,235(42.5%) had received at least one dose of the COVID-19 vaccine. The main reasons for not receiving the first dose of the vaccine included being afraid of the vaccine side effects (40.1%) or not wanting the vaccine (38.1%). The main reason for not receiving the second or third dose was still waiting for a text message for an appointment (67.1% and 73.5%, respectively). Predictors of receiving at least one dose of COVID-19 vaccine included: age, sex, residence, education and intention of vaccination. After adjusting for optimisation, the final model showed moderate discrimination (c-statistic=0.605[95%CI:0.584 to 0.624]) and good calibration (c-slope=0.912[0.758 to 1.079]).

**Interpretation:** There is an ongoing need to address vaccine acceptance and uptake among older Syrian refugees, by improving deployment planning, and raising awareness campaigns about the importance of the vaccine.

**Funding:** The study was funded by ELRHA’s Research for Health in Humanitarian Crisis (R2HC) Programme. R2HC is funded by the UK Foreign, Commonwealth and Development Office (FCDO), Wellcome, and the UK National Institute for Health Research (NIHR).

**Research in context:** *Evidence before the study:* We searched PubMed and Google Scholar for studies published up to September 29, 2022, that target COVID-19 vaccine hesitancy and uptake among older Syrian refugees in the region. We used the following combinations of keyword in our search: “COVID-19 vaccine hesitancy”, “COVID-19 vaccine uptake”, “vaccine literacy”, “vaccine acceptance”, “Syrian refugees”, “predictors”, and “education”. Previous evidence has shown that Syrian refugees have a high risk of severe morbidity and death from COVID-19 infection. Vaccine hesitancy among this vulnerable group in Lebanon is high, and there is a paucity of data available about vaccine uptake in refugee populations. In addition, older Syrian refugees have faced multiple barriers to accessing healthcare. Hence, measuring actual COVID-19 vaccine uptake and understanding the predictors influencing vaccine uptake among older Syrian refugees is critical to improving vaccination access and strategies related to COVID-19 in Lebanon.

*Added value of the study:* To our knowledge, no studies have examined predictors of COVID-19 vaccine uptake and measured the rate of vaccination among older Syrian refugees. Out of 2,906 participants, 1,235 (42.5%) received at least one dose of COVID-19 vaccine, and 806 (27.7%) received two doses of the COVID-19 vaccine. From the available data from the Ministry of Public health in Lebanon, the reported rate of vaccination among this group is low compared to the Lebanese population. In addition, this study developed a predictive model and identified five predictors of receiving at least one dose of COVID-19 vaccine among older Syrian refugees: age, sex, residence (inside or inside informal tented settlement), education and intention of vaccination.

*Implications of all the available evidence:* These findings suggest an ongoing need to address vaccine acceptance and uptake among older Syrian refugees by spreading awareness about the importance of the COVID-19 vaccine and enhancing the national system for faster vaccine coverage and response in future crises. Focusing on vaccine intention through tailored interventions and targeting hard-to-reach populations will improve vaccine uptake among Syrian refugees.

## Introduction

Vaccination against COVID-19 is crucial for mitigating the pandemic.^1^ Up until October 2022, around 68.4% of the world population has received at least one dose of the COVID-19 vaccine.^2^ Yet, in low-income countries, only 23.3% of people have received at least one dose.^2^ Although inequalities in the distribution of the vaccine between and within countries have been amply reported,^3^ disparities in vaccine hesitancy remain a main obstacle globally. For example, vaccine acceptance rates vary between 97% in Nepal and Vietnam and 13% in Iraq.^4^

Most research worldwide has focused on intention to receive the COVID-19 vaccine rather than actual vaccination status or vaccine uptake.^5-11^ According to a systematic review of international studies, predictors of COVID-19 vaccine uptake included sex, age, being a native-born rather than an immigrant or refugee, educational status, and economic status.^1^ Potential barriers to COVID-19 vaccine acceptance among refugees worldwide could be related to cultural and religious beliefs, mistrust in the health system, lack of knowledge and awareness about the importance of vaccination, lack of access to vaccination centers, and legal status (lack of required documentation for vaccine registration).^12^

Lebanon hosts approximately 1.5 million Syrian refugees, including 831,053 registered with the United Nations High Commissioner for Refugees (UNHCR).^13^ Given the country’s stretched health care system, Syrian refugees face barriers to accessing health care, which makes them more prone to under-vaccination. Moreover, they are at a higher risk for COVID-19 infection due to living in crowded conditions and lacking access to clean water and hygiene services.^12 14^ In January 2021, the Lebanese Ministry of Public Health Lebanon (MOPH) launched a National Deployment Vaccination Plan (NDVP) for COVID-19 vaccines, which aimed to vaccinate everyone on Lebanese soil, including non-citizens and refugees regardless of legal status.^14^ All Lebanese residents, regardless of nationality are allowed to register free of charge on the MOPH COVAX platform by including their personal information.^15^ The platform allows the registration of undocumented refugees or migrants without having to provide their personal ID number.^16^

Vaccine registration and administration among Syrian refugees in Lebanon remains lower than the Lebanese population.^16^ As of October 2022, 579,330 Syrian persons have registered on the platform and nearly 608,878 doses were administered to Syrian persons.^17^ Furthermore, out of all vaccines administered in Lebanon, 82.3% account for Lebanese, and only 10.7% for Syrians.^17 18^ Vaccine hesitancy among migrants and refugees in Lebanon is high, with commonly reported reasons related to concerns over vaccine safety, side effects, and registration requirements.^19^ A recently published study among older Syrian refugees in Lebanon identified a number of predictors of COVID-19 vaccine intention including sex, age, education, living outside informal tented settlements, perceiving vaccines as not safe or effective, and using social media as a source of information on COVID-19.^11^

To date, research on actual COVID-19 vaccination status among Syrian refugees in the Arab region has been very limited. Understanding the predictors that influence vaccine uptake in this vulnerable population is critical to improve vaccination programs, ensure vaccine equity, and enhance response strategies related to COVID-19 and future outbreaks. In this study, we aimed to elucidate the predictors of COVID-19 vaccine uptake among older Syrian refugees in Lebanon and understand the main reasons for not receiving the vaccine.

## Methods

### Study design and setting

This was a cross-sectional study nested within a five-wave longitudinal study that aimed to track the vulnerabilities of older Syrian Refugees residing in Lebanon to COVID-19 over time. The study followed the Transparent Reporting of a Multivariable Prediction Model for Individual Prognosis or Diagnosis (TRIPOD) reporting guideline and Strengthening the Reporting of Observational Studies in Epidemiology (STROBE) reporting guideline for prediction modelling.^20 21^

### Sampling and study population

The sampling frame included all households in Lebanon that received assistance from a humanitarian, non-governmental organization [Norwegian Refugee Council] between 2017 and 2020 and had a Syrian adult aged 50 years or older (n=17,384). All households in the sample listing were contacted and Syrian refugees aged 50 years or older were invited to participate in the study. If a household included more than one Syrian refugee aged 50 years or older, one adult was randomly chosen. Verbal consent was taken from all the participants and those aged 65 years of older were assessed for capacity to consent.^22^ Each respondent completed a computer-assisted telephone interview across five waves starting from September 2020 until March 2022 (wave 5). Figure 1 represents the flowchart of the study population for this analysis; our sample included participants who completed wave 3 (January-April 2021) and wave 5 (January-March 2022).

**Figure 1.**
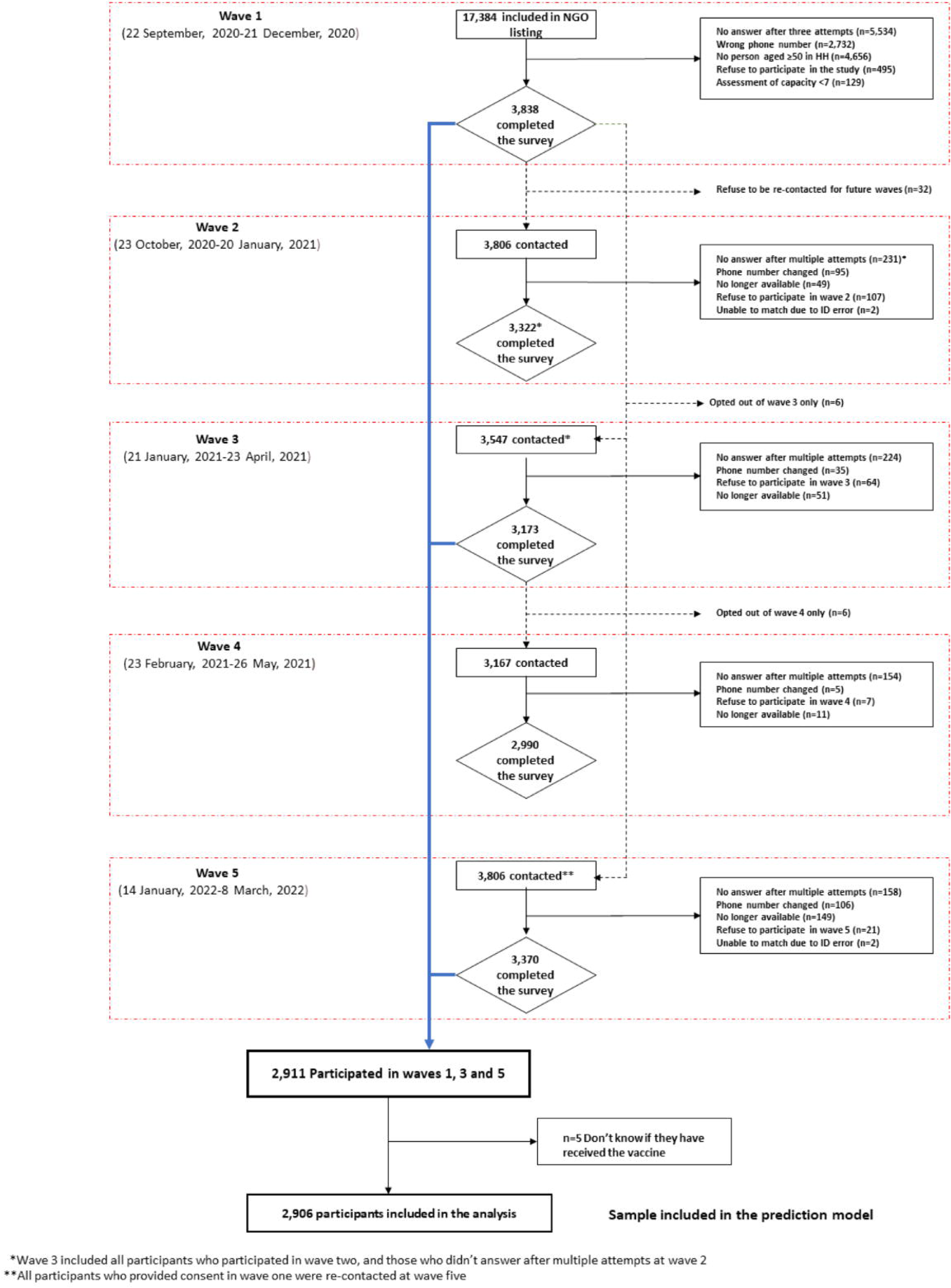
Flow diagram of Syrian refugees included in the study population.

### Data sources

The questionnaire for each wave was developed using multiple sources, including existing validated scales and contextualized questions, and was co-created by academics, humanitarian workers, government representatives, and focal points from the refugee community. The survey tool was drafted in English and then translated into Arabic. Prior to onset of data collection, the Arabic version of the questionnaire was piloted internally with data collectors and local community focal points for face validity. Several adjustments to the survey instrument were performed based on the pilot test, community consultations, and data collectors’ training to ensure contextualization and face validity. The survey was administered in Arabic over the phone by trained data collectors and data entry was undertaken using Kobo toolbox. Data monitoring was performed daily in parallel with data collection for quality assurance. More details about the development of the survey tool are available in a previously published paper.^23^ For the present analysis, data were extracted from two study waves: 1) wave 3, which was administered during the early phases of the COVID-19 vaccine roll out and included a question on whether participants intend to receive it; and 2) wave 5, which was administered months after the vaccine roll out and included questions on whether participants received one or more doses of the vaccine.

### Outcome measures

COVID-19 vaccine uptake was the primary outcome of interest. Each participant was asked the following questions in wave 5: “Have you received the COVID-19 vaccine?” and “If yes, did you receive the first/second/third dose of the vaccine?”. There were two outcomes: receiving at least one dose of COVID-19 vaccine (0=no dose; 1=at least one dose) and receiving two doses or more of COVID-19 vaccine (0=no dose; 1=two doses or more).

### Possible predictors

Based on the literature, the following possible predictors for vaccine uptake were included: 1) socio-demographic variables: age, sex, residence outside or inside informal tented settlements, education and employment status; 2) intention to receive the vaccine assessed using the following question from wave 3: “Now that a safe and effective COVID-19 vaccine has arrived to Lebanon and is offered for free, would you take it?”; 3) number of chronic disease conditions; 4) perception of vaccine safety; and 5) receipt of cash assistance.

### Missing data

The missing values were assumed to be missing at random. As they did not exceed 5% for all predictors, a complete case analysis was conducted.^24^

### Statistical analysis

Frequencies and percentages were reported for categorical variables and median and interquartile range for continuous variables. Unadjusted odds ratios (OR) along with their 95% confidence intervals (95% CI) were reported for predictors across the two outcome variables: (1) receiving at least one COVID-19 vaccine dose compared to none; (2) receiving two COVID-19 vaccine doses or more compared to none. A P-value less than 0.05 was considered statistically significant.

All the candidate predictors were categorical except age, which had a linear relationship with each outcome. A stepwise backwards multivariable logistic regression model was used where all candidate predictors were entered into the model and removed using a p-value<0.157.^25^ Multicollinearity was assessed and a variance inflation factor higher than five indicated collinearity.

The C-Statistic was used to assess the final model’s performance in terms of discrimination. It is also known as the area under the receiver operating characteristic curve (AUCROC) and ranges from 0.5 (a discriminative ability equal to chance) to 1.0 (perfect discriminative ability between those with and without the outcome). Calibration plots and slope were used to assess the model calibration where a slope of 1 and intercept of 0 (diagonal line) shows perfect calibration or agreement between observed outcomes, and a slope less than 1 indicates overfitting in the model.^26^ In addition, calibration-in-the-large (CITL) was also assessed to identify the overall difference between the observed number of events and the average predictive risk.

The final model was internally validated using bootstrap methods; where 500 bootstrap samples with replacement were used to assess the model’s optimism. The optimism adjusted estimates of C-Statistic and optimism-adjusted calibration plot were generated.^24 26-28^ Finally, bootstrap shrinkage was applied to the final apparent model giving adjusted coefficients and odds ratios. All analyses were conducted using Stata/SE 17.

### Ethical approval

This study was approved by the American University of Beirut Social and Behavioral Sciences Institutional Review Board [Reference: SBS-2020-0329]. Consent to participate was obtained verbally from all the participants.

## Results

### Characteristics of the population

Out of 3,838 Syrian refugees who completed the first wave, 3,173 participated in wave 3; of those, 2,906 participants completed wave 5 and responded to the vaccine uptake question (Figure 1). The median age of the study sample was 58 (IQR=55-64; range=52-100); slightly more than half of the sample (52.9%) were males and the majority (62.7%) lived outside tented settlements (Table 1).

**Table 1:** Characteristics of older refugees and their associations with COVID-19 vaccine uptake.

|  | Total (n=2906) |  | No dose taken (n=1671<br>(57.5%)) |  | One dose only taken<br>(n=429(14.8%)) |  | Two or more doses<br>taken (n=806 (27.7%)) |  | Two or more doses<br>taken Vs None | At least one dose Vs<br>None |
| --- | --- | --- | --- | --- | --- | --- | --- | --- | --- | --- |
|  | n | (%) | n | (%) | n | (%) | n | (%) | OR (95% CI) | OR (95%CI) |
| ***Socio-Demographics*** |  |  |  |  |  |  |  |  |  |  |
| ***Age (years) (Median (Q1-Q3))*** | 2906 (58 (55-64)) |  | 1671 (59 (55-65)) |  | 429 (57 (54-62)) |  | 806 (57 (54-62)) |  | 0.96 (0.95-0.97) | 0.96 (0.95-0.97) |
| ***Sex*** |  |  |  |  |  |  |  |  |  |  |
| Female | 1368 | (47.1) | 878 | (52.5) | 178 | (41.5) | 312 | (38.7) | 1 | 1 |
| Male | 1538 | (52.9) | 793 | (47.5) | 251 | (58.5) | 494 | (61.3) | 1.75 (1.48-2.08) | 1.68 (1.45- 1.95) |
| ***Residence*** |  |  |  |  |  |  |  |  |  |  |
| Outside informal tented settlements | 1822 | (62.7) | 1109 | (66.4) | 256 | (59.7) | 457 | (56.7) | 1 | 1 |
| Inside informal tented settlements | 1084 | (37.3) | 562 | (33.6) | 173 | (40.3) | 349 | (43.3) | 1.51 (1.27-1.79) | 1.44 (1.24-1.68) |
| ***Education*** |  |  |  |  |  |  |  |  |  |  |
| Never attended school | 1379 | (47.6) | 862 | (51.8) | 196 | (45.8) | 321 | (39.8) | 1 | 1 |
| Elementary | 762 | (26.3) | 400 | (24.0) | 117 | (27.3) | 245 | (30.4) | 1.64 (1.34-2.02) | 1.51 (1.26-1.80) |
| Preparatory and above | 757 | (26.1) | 402 | (24.2) | 115 | (26.9) | 240 | (29.8) | 1.60 (1.31-1.97) | 1.47 (1.23-1.76) |
| Missing | 8 |  | 7 |  | 1 |  | 0 |  |  |  |
| ***Health*** |  |  |  |  |  |  |  |  |  |  |
| ***Chronic conditions*** |  |  |  |  |  |  |  |  |  |  |
| At least one | 2172 | (76.4) | 1272 | (77.7) | 310 | (74.5) | 590 | (74.7) | 1 | 1 |
| None | 671 | (23.6) | 365 | (22.3) | 106 | (25.5) | 200 | (25.3) | 1.18 (0.97-1.44) | 1.18 (0.99-1.41) |
| Missing | 63 |  | 34 |  | 13 |  | 16 |  |  |  |
| ***Vaccination Perceptions*** |  |  |  |  |  |  |  |  |  |  |
| ***I think vaccines are safe*** |  |  |  |  |  |  |  |  |  |  |
| Disagree | 325 | (11.2) | 211 | (12.6) | 36 | (8.4) | 78 | (9.7) | 1 | 1 |
| Agree | 2084 | (71.7) | 1174 | (70.3) | 327 | (76.2) | 583 | (72.3) | 1.34 (1.02-1.77) | 1.43 (1.12-1.83) |
| Neither Agree nor disagree or Don't know | 497 | (17.1) | 286 | (17.1) | 66 | (15.4) | 145 | (18.0) | 1.37 (0.99-1.90) | 1.36 (1.02-1.82) |
| ***Cash assistance*** |  |  |  |  |  |  |  |  |  |  |
| No | 197 | (6.8) | 114 | (6.8) | 31 | (7.2) | 52 | (6.5) | 1 | 1 |
| Yes | 2704 | (93.2) | 1553 | (93.2) | 398 | (92.8) | 753 | (93.5) | 1.06 (0.76-1.49) | 1.02 (0.76-1.36) |
| Missing | 5 |  | 4 |  | 0 |  | 1 |  |  |  |
| ***Intention to receive vaccination*** |  |  |  |  |  |  |  |  |  |  |
| Refuse | 911 | (31.4) | 571 | (34.2) | 119 | (27.7) | 221 | (27.5) | 1 | 1 |
| Accept | 1769 | (61.0) | 966 | (57.9) | 281 | (65.5) | 522 | (65.0) | 1.40 (1.16-1.68) | 1.40 (1.18-1.64) |
| Don't know | 220 | (7.6) | 131 | (7.8) | 29 | (6.8) | 60 | (7.5) | 1.18 (0.84-1.67) | 1.14 (0.84-1.54)) |
| Missing | 6 |  | 3 |  | 0 |  | 3 |  |  |  |

### Vaccine uptake and intention to vaccinate

Out of 2,906 participants, 1,671 (57.5%) reported not receiving any dose of COVID-19 vaccine in wave 5; 429 (14.8%) reported only one dose received; and 806 (27.7%) reported two or more doses received (Table 1 and Figure 2). The main reasons for not receiving the first dose of the vaccine included being afraid of the vaccine side effects (670[40.1%]), followed by not wanting the vaccine (637[38.1%]) and still waiting to register or to be called for an appointment (348[20.8%]). In addition, the main reasons for not receiving the second dose of the vaccine were that the participants were waiting for a text message for an appointment after having registered (288[67.1%]) or reported being afraid of the vaccine’ side effects or not wanting to receive it (59[13.7%]). Moreover, among those who had received a second dose, but not a third, 73.5% (n=573) were waiting for a text message to schedule an appointment, and 10% (n=78) didn’t want to receive the vaccine or considered that two doses were enough (Figure 2).

**Figure 2.**
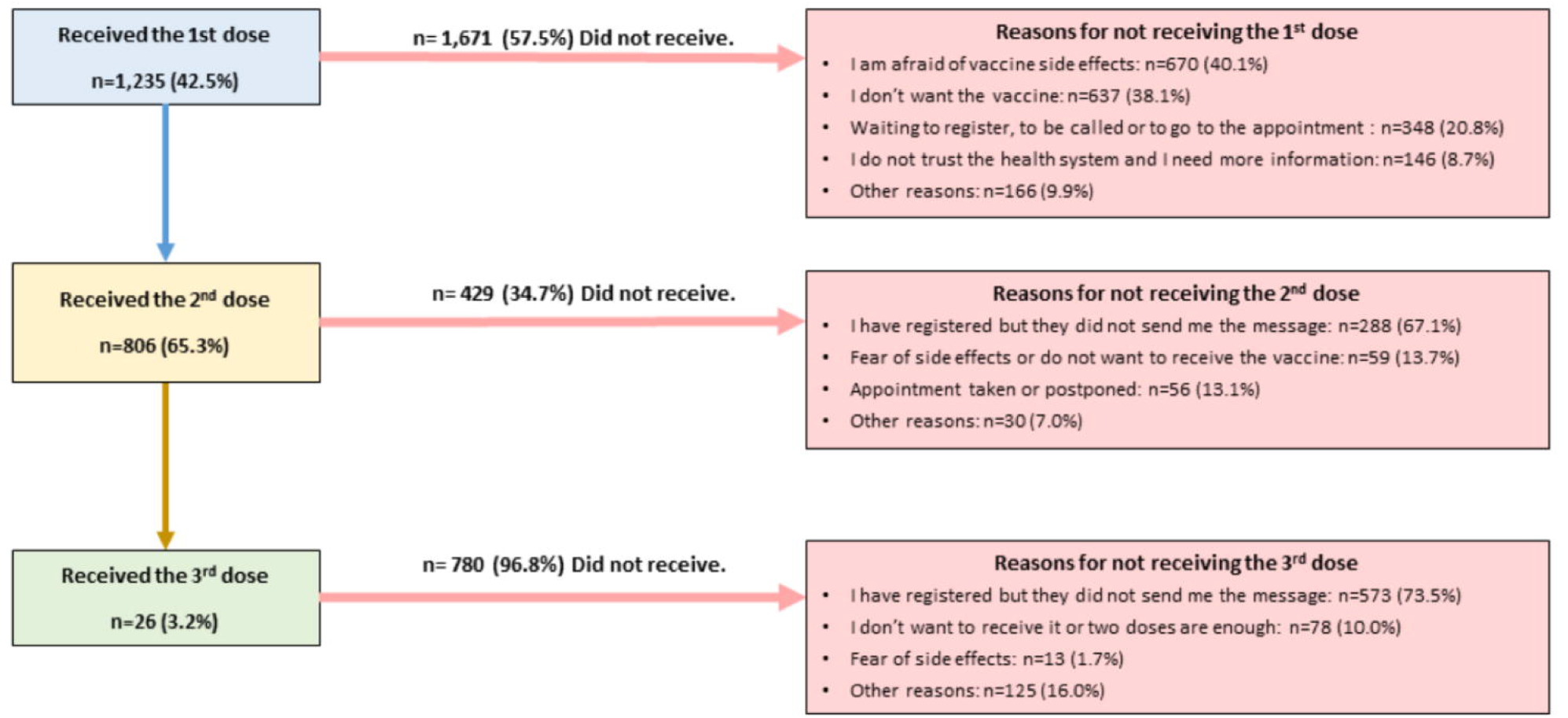
Vaccination prevalence and reasons for not receiving Dose 1, Dose 2, and Dose 3 of COVID-19 vaccine. Footnote: *Includes those who were unable to access the vaccine center, sickness, get infected by COVID-19, afraid from needle, etc.

### Bivariate predictors of COVID-19 vaccine uptake

The unadjusted analysis showed that older age (OR=0.96[95%CI:0.95 to 0.97]) is associated with decreased odds of receiving at least one dose of COVID-19 vaccine. Furthermore, participants who were more likely to receive at least one dose of the vaccine in wave 5 were male compared to female (OR: 1.68[95%CI:1.45 to 1.95]), lived inside informal tented settlements (OR=1.44[95%CI:1.24 to 1.68]) compared to living outside, had elementary (OR=1.51[95%CI:1.26 to1.80]) and preparatory education or above (OR=1.47[95%CI:1.23 to 1.76]) compared to never having attended school, agreed with the statement that “vaccines are safe” (OR=1.43[95%CI:1.12 to 1.83]) compared to disagreeing with it, and had reported intention to receive the vaccine in wave 3 (OR=1.40[95%CI:1.18 to 1.64]) compared to those who reported no intention to receive it. The characteristics of Syrian refugees who were likely to take at least one dose of the vaccine were similar to those who were likely to take two or more doses of the vaccine (Table 1).

### Predictors and model performance

The final model identified five predictors of receiving at least one dose of COVID-19 vaccine in wave 5 which included: age, sex, residence, education, and earlier intention to receive the vaccine (Table 2). Figure 3 presents the calibration plot adjusted for optimism, whereas the calibration plot of the apparent model before correction for optimism is shown in the appendix (Figure 1; p 2). Thereby, after adjustment for optimism, the final model had a C-statistic of 0.605[95%CI:0.584 to 0.624] which indicates a moderate discriminative ability, as well as a calibration slope of 0.912[95%CI:0.758 to 1.079], the brier score of 3.3, and CITL of 0.001[95%CI: −0.074 to 0.073] (Table 2 and Figure 3). The odds ratios along with their coefficients have been adjusted using bootstrap shrinkage and are presented in Table 2.

**Table 2:**
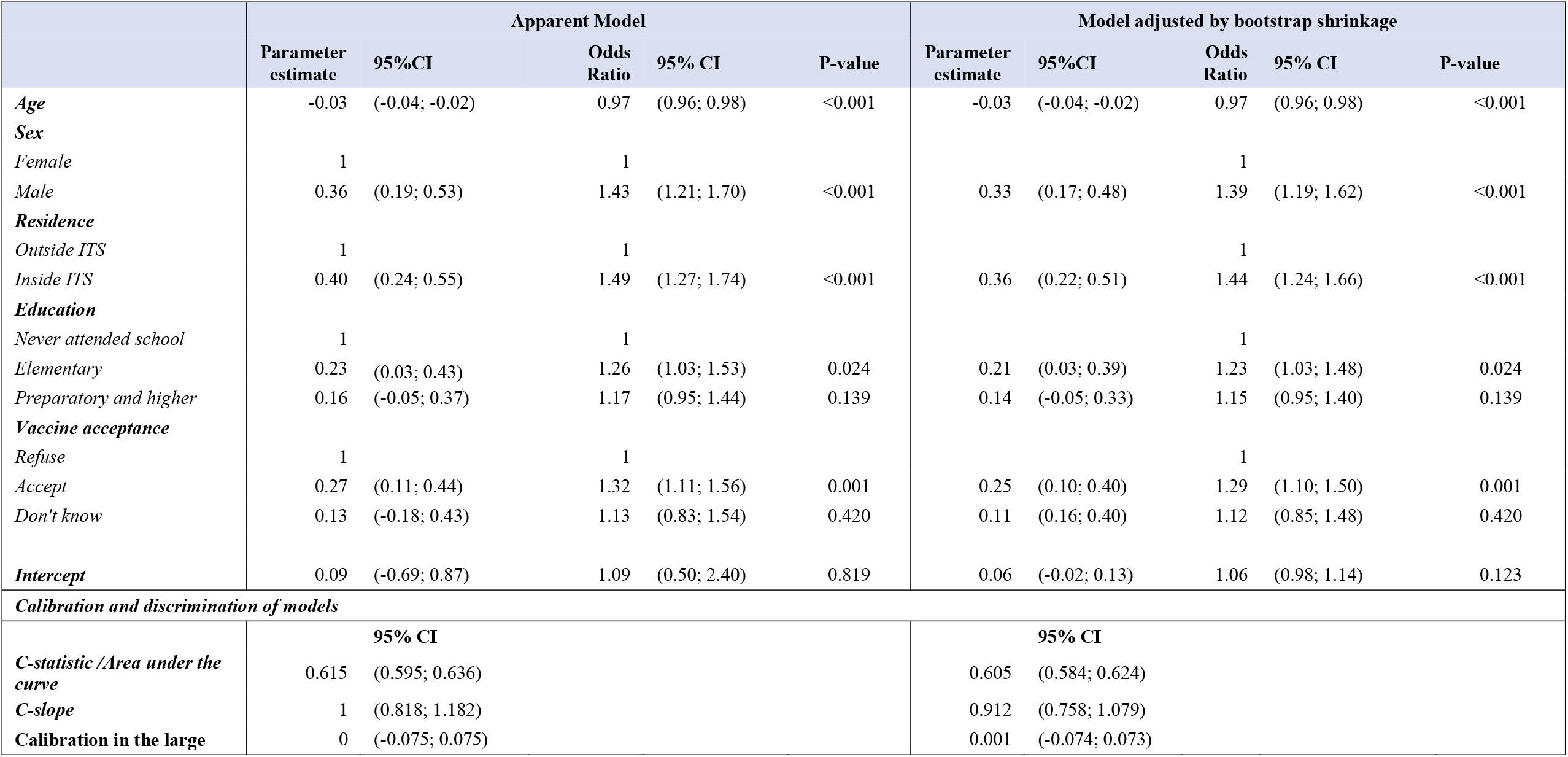
Multivariable model for predicting COVID-19 vaccine uptake of at least one dose.

**Figure 3:**
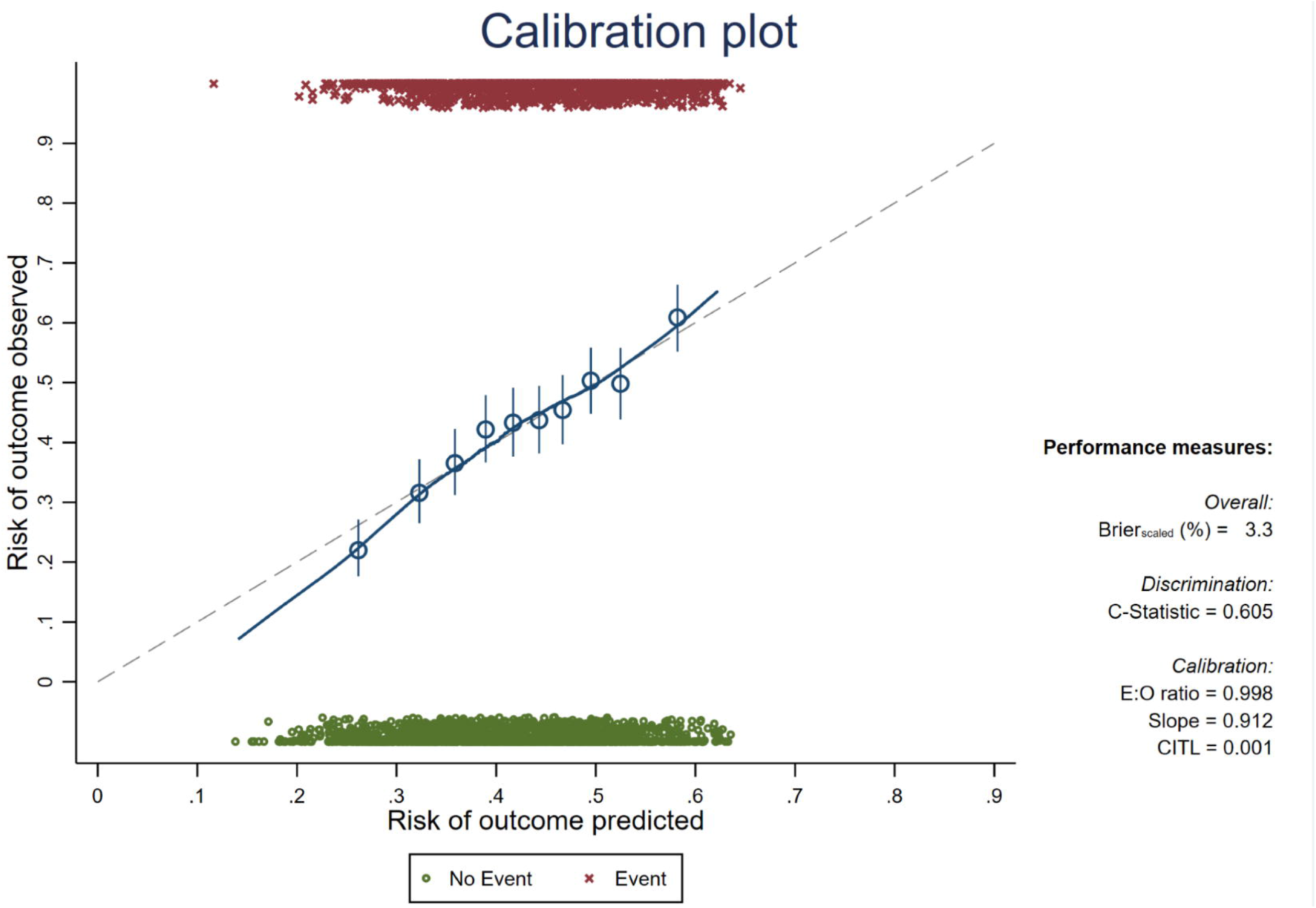
Model performance in the optimized-adjusted model

A second predictive model examining the predictors of receiving at least two doses of the vaccine compared to none is presented in the appendix (table 1; p 3). It identified the same five predictors as the model predicting at least one dose of the vaccine. The final model also had an optimized adjusted C-statistic of 0.611[95%CI:0.588 to 0.635] and a calibration slope of 0.905[95%CI:0.750 to 1.080].

The direction of predictors’ coefficients with the outcome were all in the expected direction; being a male compared to female, living inside informal tented settlements compared to living outside, having higher education, and reporting intention to receive the vaccine in wave 3 had positive coefficients and therefore higher probability of receiving at least one dose of the COVID-19 vaccine in wave 5. Only age had a negative coefficient, meaning that those with higher age had a lower likelihood of receiving the vaccine.

To illustrate, the predicted probability of receiving at least one dose of COVID-19 vaccine, for a Syrian refugee who is a female, aged 85 years, never attended school, residing outside informal tended settlements and did not intend to receive the vaccine was 7.6%. However, for a Syrian refugee who is a male, aged 50 years, having an elementary education, residing inside informal tended settlements and intended to receive the vaccine, the predicted probability was 42.8%.

## Discussion

This study showed that more than half of older Syrian refugees in Lebanon had not received any dose of the COVID-19 vaccine by wave 5; 42.5% of older Syrian refugees had received at least one dose. It also identified five predictors of receiving at least one dose of COVID-19 vaccine among older Syrian refugees: age, sex, residency (inside or outside informal tented settlements), education, and having reported the intention to receive the vaccine. The predictive model had a moderate discrimination ability and good calibration. The primary reasons for not receiving the vaccine included being afraid of the vaccine side effects, not wanting to receive the vaccine, or waiting to be invited to receive the vaccine after registering on the platform. The main reason for not receiving the second or third dose of the vaccine was mainly waiting for an appointment after having registered.

## Results in context

Given the magnitude of the COVID-19 pandemic, the level of vaccination among this vulnerable group is considerably low, particularly in comparison to the suggested average threshold to reach COVID-19 herd immunity is approximately 67%.^29^ The observed prevalence of vaccine uptake among older Syrian refugees (42.5%) could not be compared to the host population because the national prevalence by nationality is not available.^17^ Despite the efforts exerted by international organizations and the Ministry of Public Health in Lebanon to ease the vaccine registration process among Syrian refugees,^16^ our results show that vaccination registration and uptake among Syrian refugees in Lebanon are low (more details on available data can be found in the appendix (table 2; p 4)).^17^ Hence, this indicates an urgent need for public health interventions that highlight the importance of the vaccine. The measured prevalence of vaccine uptake in this study is higher than that of a study conducted among refugees living in Australia, where only 12% reported to be vaccinated.^6^

The main reasons for not receiving the vaccine that emerged from this study included not wanting the vaccine or being afraid of its side effects. Although studies that target vaccine acceptance among Syrian refugees in the Middle East are very limited, these findings correspond with other studies on migrants in the UK and on the general population in Europe and the US.^8 30^ Other reasons for not taking the vaccine reported in the literature include concerns about the effectiveness of the vaccine, religious reasons, limited knowledge about the importance of the vaccine, and the belief that COVID-19 is not a serious infection.^1^ To counter misinformation about COVID-19 vaccine and improve trust, community-based interventions need to be addressed to high-risk populations such as Syrian refugees. Moreover, it may be beneficial for COVID-19 information to be shared by healthcare professionals and local and religious leaders rather than only receiving information about COVID-19 through social media.^7^

The main reason for not receiving the second or third doses of the vaccine was that the participants were still waiting for an appointment after registering. The Lebanese government must prioritize this population, that is eager to receive additional doses but who are delayed by the system, and collaborate with humanitarian agencies to strengthen the national immunization system by reducing the long waiting time between registration and appointments. Improving COVID-19 vaccination system strategies are crucial to prevent recurring future outbreaks in Lebanon, and can strengthen the response to other emerging outbreaks and future health threats.

Our study showed that males were more likely to receive at least one dose of COVID-19 vaccine compared to females. Although research about vaccine uptake is scarce, this finding is similar to the results of studies conducted in Kuwait and in the US, which showed that males were less likely to report vaccine hesitancy and tend to accept to take the vaccine more than women.^7 9^ Further research should aim to understand the reasons for this difference between the sexes that is embedded in how unequal gender roles operate in the particular context of displacement. Because older Syrian refugee women engage less than men outside the privacy of the home, they may believe that they have low risk and thus do not need to receive the vaccine. Through a gendered lens, humanitarian organizations could better understand the link between gender and vaccine uptake and, thus, would be able to design tailored and more effective awareness programs.

Another predictor for vaccine uptake that emerged from this study was younger age, which is in contrast to a study conducted on the general population in the US showing that vaccine uptake increases with age.^10^ Such conflicting results could be due to regional differences in population perceptions about vaccination, which varies across age groups. Nonetheless, this is an alarming finding in light of evidence that age is a biological risk for more severe COVID-19 illness. It is therefore critical that humanitarian organizations focus their efforts on older age groups to understand whether their low vaccine uptake is due to hesitancy or lack of trust on the one hand, or limited ability to use technology to register on the other hand.

As expected, older Syrian refugees with higher levels of education were more likely to receive the COVID-19 vaccine compared to those who never attended school. Although most studies in the literature examine the relationship between vaccine hesitancy (not uptake) and education, this finding corresponds with previous studies conducted in the US on health care personnel and the general population.^31 32^ Furthermore, vaccine literacy can increase vaccine uptake as it encompasses knowledge about the importance of the vaccine.^33^

Older Syrian refugees living inside informal tented settlements had a higher likelihood of receiving the vaccine than those in residential areas. Refugees in enclosed geographic areas like camps are easier to locate than those living in residential areas,^34 35^ which makes them easier to reach by humanitarian organizations through mobile clinics or vaccination campaigns. To prevent inequities, humanitarian organizations need to expand their programs to reach all refugees through registration centers, community centers, community-based volunteers, mobile health clinics, and municipalities.

Additional predictors for vaccine uptake identified in the literature, but not present in our model, included self-perceived vulnerability to COVID-19 and information about COVID-19 vaccines.^1^ These missing predictors may have improved the discrimination of our study’s model. Hence, additional studies in the Middle East are crucial to further understand the characteristics of the communities at risk of having low vaccination rate, to design tailored vaccination programs related to COVID-19, and ultimately improve the rate of vaccine uptake, specifically among vulnerable populations.

## Strengths and limitations

To our knowledge, this study is the first of its kind in examining predictors of COVID-19 vaccine uptake and measuring the rate of vaccination among older Syrian refugees. In addition, it is one of the largest studies on older Syrian refugees with a response rate higher than 85%.

This study is subject to several limitations. The study outcome was self-reported and may be prone to information bias. Hence, future studies should aim to have more objective outcomes. Furthermore, the study sample is representative of beneficiaries from a single humanitarian organization but not all Syrian refugees residing in Lebanon. Hence, national prevalence of vaccine uptake among older Syrian refugees cannot be measured and our findings cannot be generalized.

## Conclusion

As COVID-19 vaccine is widely available in Lebanon, our findings confirm the ongoing need to address vaccine acceptance and uptake among older Syrian refugees. Access to vaccination by the most vulnerable populations such as Syrian refugees, is crucial to curtail the spread of COVID-19 virus and other outbreaks. Lebanon, a country already overwhelmed by its economic and political crises requires further strengthening to improve deployment planning, raise awareness about the importance of vaccination, and enhance the national system for a faster response in future health crises.

## Data Availability

The anonymised data can be requested upon reasonable request from NRC and AUB.

## Acknowledgments

The authors would like to thank Mrs. Noura Salibi and Miss Maria El Haddad for their support and expertise throughout all aspects of the research study, including data monitoring, cleaning and analysis. The authors would also like to acknowledge B.O.T (Bridge. Outsource. Transform) for their assistance in collecting the data required for the success of this study and the study participants for their participation.

## Authors contributions

SA, HG, and SM conceptualized the study and the survey design. SA, SM, BA and TK contributed to data collection and analysis. BA and TK contributed to the literature search and wrote the first draft of the manuscript; the following drafts were reviewed and revised by SM and SA. The underlying survey data were verified by SM and BA. SM supervised BA and TK throughout the project. ZR was involved in the administration and implementation of the study. ZR, SAn, and MA contributed in the interpretation of the results. All authors have reviewed and approved the final version of the paper.

## Data sharing statement

The anonymised data can be requested upon reasonable request from NRC and AUB.

## Online Appendix Content

Figure 1: Model performance for the apparent model

Table 1: Multivariable model for predicting the COVID-19 vaccine uptake of two doses and more

Table 2: Available data about COVID-19 vaccine registration and uptake among Lebanese and Syrians residing in Lebanon, as of September 14, 2022

