## Appendix for "Predictors and barriers to vaccination among older Syrian refugees in Lebanon: a multi-wave survey"

**Online Appendix Content**

### **Figure 1: Model performance for the apparent model**

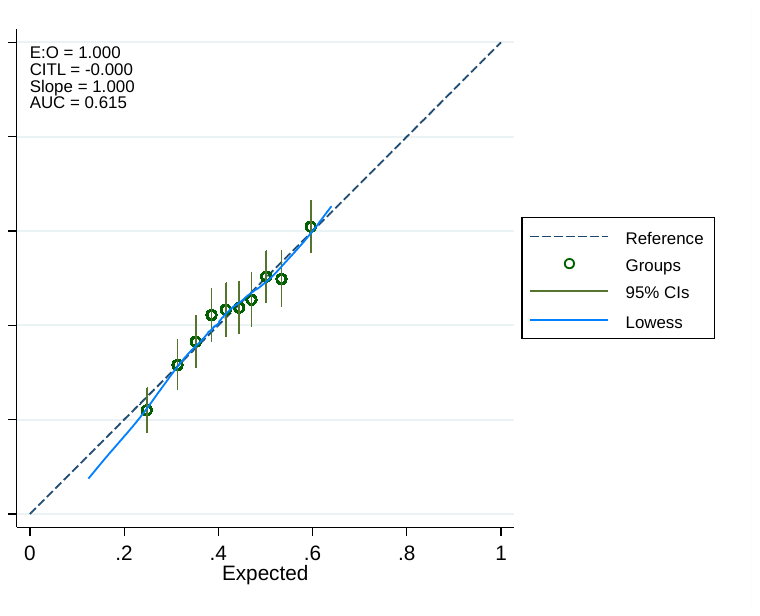

### **Table 1: Multivariable model for predicting the COVID-19 vaccine uptake of two doses and more**

|  | **Apparent Model** | | | | | **Model adjusted by bootstrap shrinkage** | | | | |
| --- | --- | --- | --- | --- | --- | --- | --- | --- | --- | --- |
|  | **Parameter estimate** | **(95%CI)** | **Odds Ratio** | **(95% CI)** | **P-value** | **Parameter estimate** | **(95%CI)** | **Odds Ratio** | **(95% CI)** | **P-value** |
| Age | -0.03 | (-0.04; -0.02) | 0.97 | (0.96; 0.98) | <0.001 | -0.03 | (-0.04; -0.01) | 0.97 | (0.96; 0.98) | <0.001 |
| Sex |  |  |  |  |  |  |  |  |  |  |
| Female | 1 |  | 1 |  |  |  |  | 1 |  |  |
| Male | 0.35 | (0.16; 0.54) | 1.42 | (1.17; 1.72) | <0.001 | 0.32 | (0.14; 0.49) | 1.37 | (1.15; 0.65) | <0.001 |
| Residence |  |  |  |  |  |  |  |  |  |  |
| Outside ITS | 1 |  | 1 |  |  |  |  | 1 |  |  |
| Intside ITS | 0.44 | (0.26; 0.62) | 1.55 | (1.30; 1.86) | <0.001 | 0.40 | (0.23; 0.56) | 1.48 | (1.26; 1.75) | <0.001 |
| Education |  |  |  |  |  |  |  |  |  |  |
| Never attended school | 1 |  | 1 |  |  |  |  | 1 |  |  |
| Elementary | 0.33 | (0.10; 0.55) | 1.39 | (1.11; 1.74) | 0.004 | 0.30 | (0.09; 0.50) | 1.34 | (1.10; 1.65) | 0.004 |
| Preparatory and higher | 0.26 | (0.02; 0.50) | 1.30 | (1.02; 1.65) | 0.030 | 0.24 | (0.02; 0.45) | 1.27 | (1.02; 1.57) | 0.030 |
| Vaccine acceptance |  |  |  |  |  |  |  |  |  |  |
| Refuse | 1 |  | 1 |  |  |  |  | 1 |  |  |
| Accept | 0.27 | (0.07; 0.46) | 1.31 | (1.08; 1.59) | 0.006 | 0.24 | (0.07; 0.42) | 1.27 | (1.07; 1.52) | 0.006 |
| Don't know | 0.17 | (-0.17; 0.52) | 1.19 | (0.84; 1.69) | 0.328 | 0.16 | (-0.16; 0.47) | 1.17 | (0.85; 1.61) | 0.328 |
| Intercept | -0.46 | (-1.38; 0.45) | 0.63 | (0.25; 1.57) | 0.323 | 0.48 | (-0.57; -0.40) | 0.62 | (0.56; 0.67) | <0.001 |
| **Calibration and discrimination of models** | | |  |  |  |  |  |  |  |  |
|  |  | **95% CI** |  |  |  |  | **95% CI** |  |  |  |
| C-statistic /Area under the curve | 0.622 | (0.599; 0.646) |  |  |  | 0.611 | (0.588; 0.635) |  |  |  |
| C-slope | 1 | (0.806; 1.194) |  |  |  | 0.905 | (0.750; 1.080) |  |  |  |
| Calibration in the large | 0 | (-0.086; 0.086) |  |  |  | 0.002 | (-0.081; 0.089) |  |  |  |

### **Table 2: Available data about COVID-19 vaccine registration and uptake among Lebanese and Syrians residing in Lebanon, as of September 14, 2022**

|  | **Lebanese** | **Syrians** |
| --- | --- | --- |
| Residence numbers^1 2^ | 3,864,315 | 1,500,000 |
| Registration numbers^3^ | 2,874,541 | 593,140 |
| Numbers of COVID-19 vaccine taken doses^3^ | 4,546,556 | 597,096 |
